# Mortality and risk factors among US Black, Hispanic, and White patients with COVID-19

**DOI:** 10.1101/2020.09.08.20190686

**Authors:** Tomi Jun, Sharon Nirenberg, Patricia Kovatch, Kuan-lin Huang

## Abstract

**Background:** Little is known about risk factors for COVID-19 outcomes, particularly across diverse racial and ethnic populations in the United States.

**Methods:** In this prospective cohort study, we followed 3,086 COVID-19 patients hospitalized on or before April 13, 2020 within an academic health system in New York (The Mount Sinai Health System) until June 2, 2020. Multivariable logistic regression was used to evaluate demographic, clinical, and laboratory factors as independent predictors of in-hospital mortality. The analysis was stratified by self-reported race and ethnicity.

**Findings:** A total of 3,086 COVID-19 patients were hospitalized, of whom 680 were excluded (78 due to missing race or ethnicity data, 144 were Asian, and 458 were of other unspecified race/ethnicity). Of the 2,406 patients included, 892 (37.1%) were Hispanic, 825 (34.3%) were black, and 689 (28.6%) were white. Black and Hispanic patients were younger than White patients (median age 67 and 63 vs. 73, p<0.001 for both), and they had different comorbidity profiles. Older age and baseline hypoxia were associated with increased mortality across all races. There were suggestive but non-significant interactions between Black race and diabetes (p=0.09), and obesity (p=0.10). Among inflammatory markers associated with COVID-19 mortality, there was a significant interaction between Black race and interleukin-1-beta (p=0.04), and a suggestive interactions between Hispanic ethnicity and procalcitonin (p=0.07) and interleukin-8 (p=0.09).

**Interpretation:** In this large, racially and ethnically diverse cohort of COVID-19 patients in New York City, we identified similarities and important differences across racial and ethnic groups in risk factors for in-hospital mortality.

**Funding:** Icahn School of Medicine at Mount Sinai, New York, NY.

## Introduction

Reports from the United States, the United Kingdom, and Brazil have highlighted racial disparities in the COVID-19 pandemic.^1–4^ National studies from the UK and Brazil have found race to be an independent predictor of death.^3,4^ In the United States, Black and Hispanic individuals have disproportionately high rates of infection and hospitalization, and mortality.^1^’^2,5,6^ These disparities have been attributed to greater representation of Black and Hispanic persons in essential services, and a higher burden of comorbidities in minority communities, among others.

While Black and Hispanic individuals in the US have been disproportionately affected by the pandemic, the majority of published studies investigating COVID-19 mortality risk factors have been in cohorts of individuals with predominantly European or Asian ancestry.^7–11^ Only one US study has directly examined mortality risk factors and their effect sizes in Black compared to White individuals,^1^ and to our knowledge, no studies have examined this question among Hispanic individuals in the US.

The objective of this study was to identify race/ethnicity-specific associations of risk factors for COVID-19 mortality; that is, factors that may disproportionately increase the risk of death within one racial/ethnic subgroup compared to others. Although prior studies have included multivariable regression models adjusting for race/ethnicity, they have not explored the possibility of interactions between race/ethnicity and other predictors. Models without interaction terms assume that predictors have the same effect on mortality risks across racial/ethnic groups, an assumption that has not been tested. We sought to identify race/ethnic-specific associations using a diverse cohort of White, Black, and Hispanic COVID-19 patients admitted to a single health system in New York during the initial pandemic surge.

## Methods

### Study setting

The study was conducted within the Mount Sinai Health System, which is an academic healthcare system comprising 8 hospitals and more than 410 ambulatory practice locations in the New York metropolitan area. This analysis involves patients who presented to five hospitals: The Mount Sinai Hospital (1,134 beds), Mount Sinai West (514 beds), and Mount Sinai Morningside (495 beds) in Manhattan; Mount Sinai Brooklyn (212 beds); and Mount Sinai Queens (235 beds).

### Data sources

Data were captured by the Epic electronic health record (Epic Systems, Verona, WI), and directly extracted from Epic’s Clarity and Caboodle servers. This de-identified dataset was developed and released by the Mount Sinai Data Warehouse (MSDW) team, with the goal of encompassing all COVID-19 related patient encounters within the Mount Sinai system, accompanied by selected demographics, comorbidities, vital signs, medications, and lab values. As part of de-identification, all patients over the age of 89 had their age set to 90.

This study utilized de-identified data extracted from the electronic health record and as such was considered non-human subject research. Therefore, this study was granted an exemption from the Mount Sinai IRB review and approval process.

### Patient population and definitions

The MSDW dataset captured any patient encounters at a Mount Sinai facility with any of the following: a COVID-19 related encounter diagnosis, a COVID-19 related visit type, a SARS-CoV-2 lab order, a SARS-CoV-2 lab result, or a SARS-CoV-2 lab test result from the New York State Department of Health’s Wadsworth laboratory. For this study, we identified patient encounters on or before April 13^th^, 2020 and followed their mortality outcomes through June 2^nd^, 2020.

Our analysis was limited to adults over 18 years old who were hospitalized for COVID-19 through a Mount Sinai emergency department. Self-reported race and ethnicity were classified into 3 mutually exclusive categories: Non-Hispanic White (White), Non-Hispanic Black (Black), and Hispanic (**Supplemental Table 1**). COVID-19 positivity was determined by a positive or presumptive positive result from a nucleic acid-based test for SARS-CoV-2 in nasopharyngeal or oropharyngeal swab specimens. Initial vital signs were the first documented vital signs for the encounter. Hypoxia was defined as an oxygen saturation less than 92%. We defined initial labs as the first lab value within 24 hours of the start of the encounter.

### Logistic regression

The primary outcome was in-hospital mortality. Univariable and multivariable logistic regression were used to identify factors associated with death. Race/ethnicity-specific risk factors were identified by 1) constructing stratified models for each racial/ethnic category, and 2) constructing models including interaction terms between race/ethnicity and other covariates. Separate interaction models were created to test the interactions of either Hispanic ethnicity or Black race with other covariates. Interactions were compared against White race as the reference group.

We analyzed demographic factors, comorbidities, initial vital signs, baseline lab values, and treatment facility site (Manhattan vs. Brooklyn/Queens) as covariates. There was minimal clustering of outcomes by treatment site (ICC (ρ) = 0.026), and this was modeled as a fixed effect. Covariates were chosen *a priori* based on prior reports. We report the odds ratios derived from the coefficients of each model, along with the Wald-type confidence interval, and p-values.

Laboratory value analysis

Markers of inflammation, such as C-reactive protein (CRP), ferritin, and d-dimer, have been proposed as being correlated with COVID-19 severity. However, the missingness of these lab values varied across sites. Given the possibility of confounding by indication (if providers ordered these labs in more acutely ill patients), we limited the analyses involving lab tests to those obtained at the largest site (The Mount Sinai Hospital) and which had less than 15% missing values at that site. The cytokines interleukin-1-beta, interleukin-6, interleukin-8, and tumor necrosis factor-alpha were exempted from this threshold because they were obtained on a subset of COVID-19 patients in the context of a study with broad inclusion criteria.^12,13^

To test the associations of these lab values with mortality, we tested each lab test in race/ethnicity-stratified multivariable logistic regression models adjusting for age, sex, and hypoxia. The number of covariates in the model was limited due to the reduced sample size. Labs were standardized to a mean of 0 and a standard deviation of 1 prior to regression analysis.

### Statistical analysis

Patient characteristics and baseline vitals and labs were described using medians and ranges for continuous variables, and proportions for categorical variables. Continuous variables were compared using the Wilcoxon rank-sum test, and categorical variables were compared using Fisher’s exact test. All statistical analyses and data visualizations were carried out using R 4.0.0 (The R foundation, Vienna, Austria), along with the *tidyverse, ggpubr, forestplot*, and *Hmisc* packages. Statistical significance was defined as p<0.05.

## Results

### Study population

There were 3,086 adult patients with COVID-19-related hospitalizations on or before April 13^th^, 2020. Of these, we excluded 78 patients with missing race or ethnicity data, 144 Asian patients, and 458 patients with other or unspecified race/ethnicity (**Figure** 1). The remaining 2,406 patients were 37.1% Hispanic, 34.3% Black, and 28.6% White.

**Figure 1:**
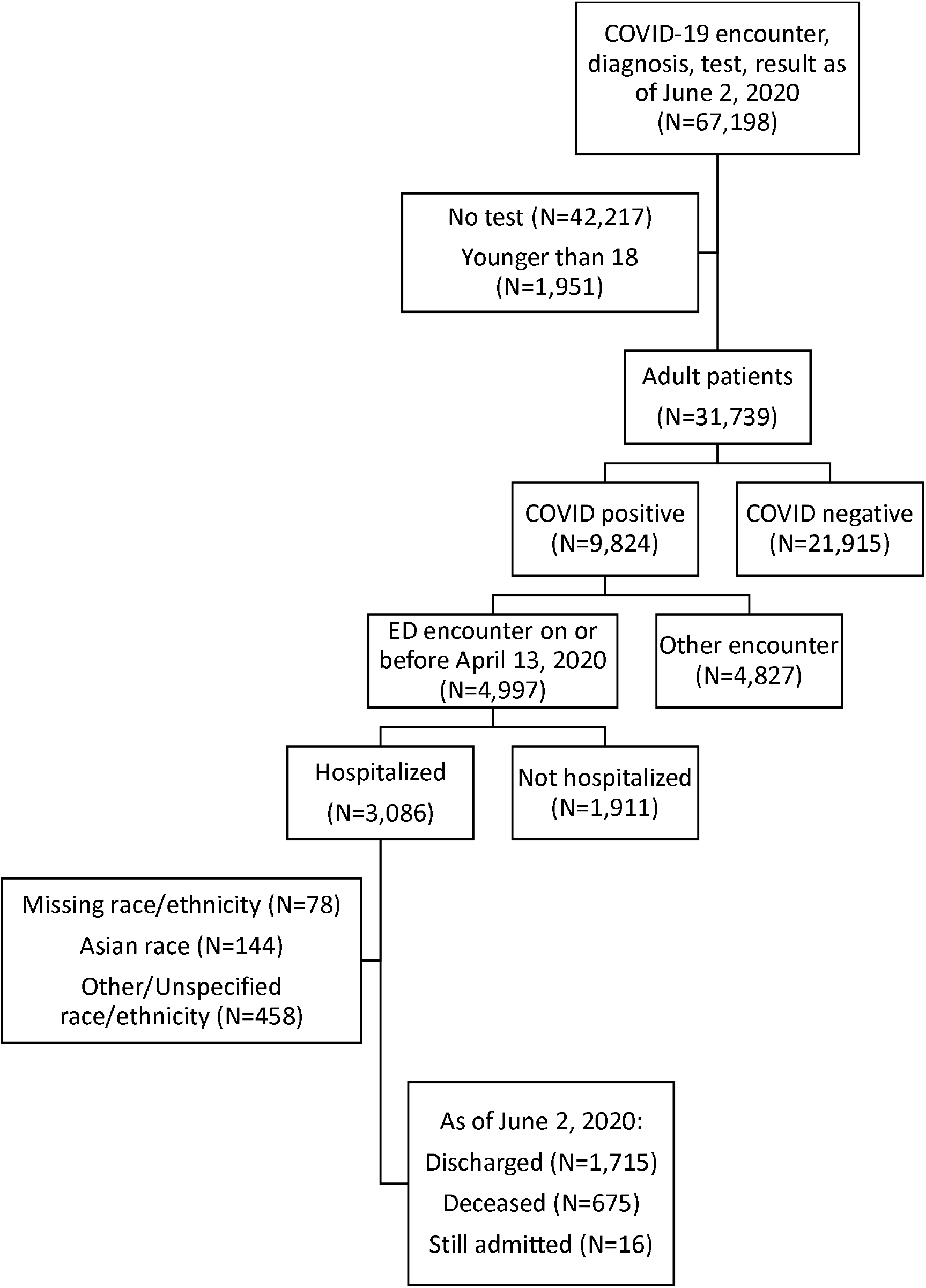
Flow diagram of included patients

Compared to White patients, Black and Hispanic patients were younger (median age 67 and 63 vs. 73, p<0.001 for both) (**Table** 1). Black patients were more likely to have hypertension (40.6% vs 31.8%, p<0.001), diabetes (26.8% vs. 17.4%, p<0.001), and chronic kidney disease (16.1% vs. 8%, p<0.001) than White patients. Hispanic patients were more likely to have diabetes (27.4% vs. 17.4%, p<0.001), chronic kidney disease (13% vs. 8%, p=0.001), and chronic liver disease (4.3% vs. 1.6%, p=0.003), compared to White patients. White patients were more likely to have coronary artery disease (17.1% vs. 12.2% and 11.3%, p=0.008 vs. Black, p=0.001 vs. Hispanic) and atrial fibrillation (12.3% vs. 5% and 4.5%, p<0.001 for both) than Black or Hispanic patients.

**Table 1:** Baseline demographic and clinical characteristics, by race/ethnicity

|  | **White**<br>**(N=689)** | **Black**<br>**(N=825)** | **Hispanic**<br>**(N=892)** |
| --- | --- | --- | --- |
| Age (yrs) | 73 (62 - 84) | 67 (58 - 76)† | 63 (50 - 74)† |
| Mount Sinai Brooklyn (MSB) | 156 (22.6%) | 258 (31.3%)† | 17 (1.9%)† |
| Mount Sinai Queens (MSQ) | 138 (20%) | 61 (7.4%)† | 271 (30.4%)† |
| Mount Sinai Morningside (MSSL) | 51 (7.4%) | 199 (24.1%)† | 216 (24.2%)† |
| Mount Sinai West (MSW) | 120 (17.4%) | 65 (7.9%)† | 100 (11.2%)† |
| The Mount Sinai Hospital (MSH) | 224 (32.5%) | 242 (29.3%) | 288 (32.3%) |
| Current smoker* | 22 (3.2%) | 43 (5.2%) | 25 (2.8%) |
| Former smoker* | 147 (21.3%) | 187 (22.7%) | 190 (21.3%) |
| Never smoker* | 375 (54.4%) | 437 (53%) | 478 (53.6%) |
| Hypertension | 219 (31.8%) | 335 (40.6%)† | 318 (35.7%) |
| Diabetes | 120 (17.4%) | 221 (26.8%)† | 244 (27.4%)† |
| Coronary artery disease | 118 (17.1%) | 101 (12.2%)† | 101 (11.3%)† |
| Heart failure | 59 (8.6%) | 71 (8.6%) | 60 (6.7%) |
| Atrial fibrillation | 85 (12.3%) | 41 (5%)† | 40 (4.5%)† |
| Chronic kidney disease | 55 (8%) | 133 (16.1%)† | 116 (13%)† |
| COPD/asthma | 60 (8.7%) | 75 (9.1%) | 84 (9.4%) |
| Obesity | 50 (7.3%) | 75 (9.1%) | 80 (9%) |
| Cancer | 52 (7.5%) | 63 (7.6%) | 57 (6.4%) |
| Chronic liver disease | 11 (1.6%) | 24 (2.9%) | 38 (4.3%)† |
| Obstructive sleep apnea | 15 (2.2%) | 18 (2.2%) | 21 (2.4%) |
| HIV | 12 (1.7%) | 23 (2.8%) | 13 (1.5%) |
| Temperature (°F) | 98.7 (98 - 99.9) | 98.9 (98.1 - 100.1)† | 99 (98.3 - 100.4)† |
| Heart rate (bpm) | 91 (79 - 106) | 97 (85 - 110)† | 99 (86 - 113)† |
| Systolic blood pressure (mmHg) | 129 (114 - 146) | 131 (116 - 148) | 128 (115 - 144) |
| Respiratory rate (bpm) | 20 (18 - 22) | 20 (18 - 22) | 20 (18 - 22) |
| Oxygen sat. <92% | 184 (26.7%) | 149 (18.1%)† | 257 (28.8%) |

Values represent count (%) or median (IQR) for categorical and continuous variables, respectively. †: p<0.05 compared to White. \*: 22% missing values

**Table 2:**
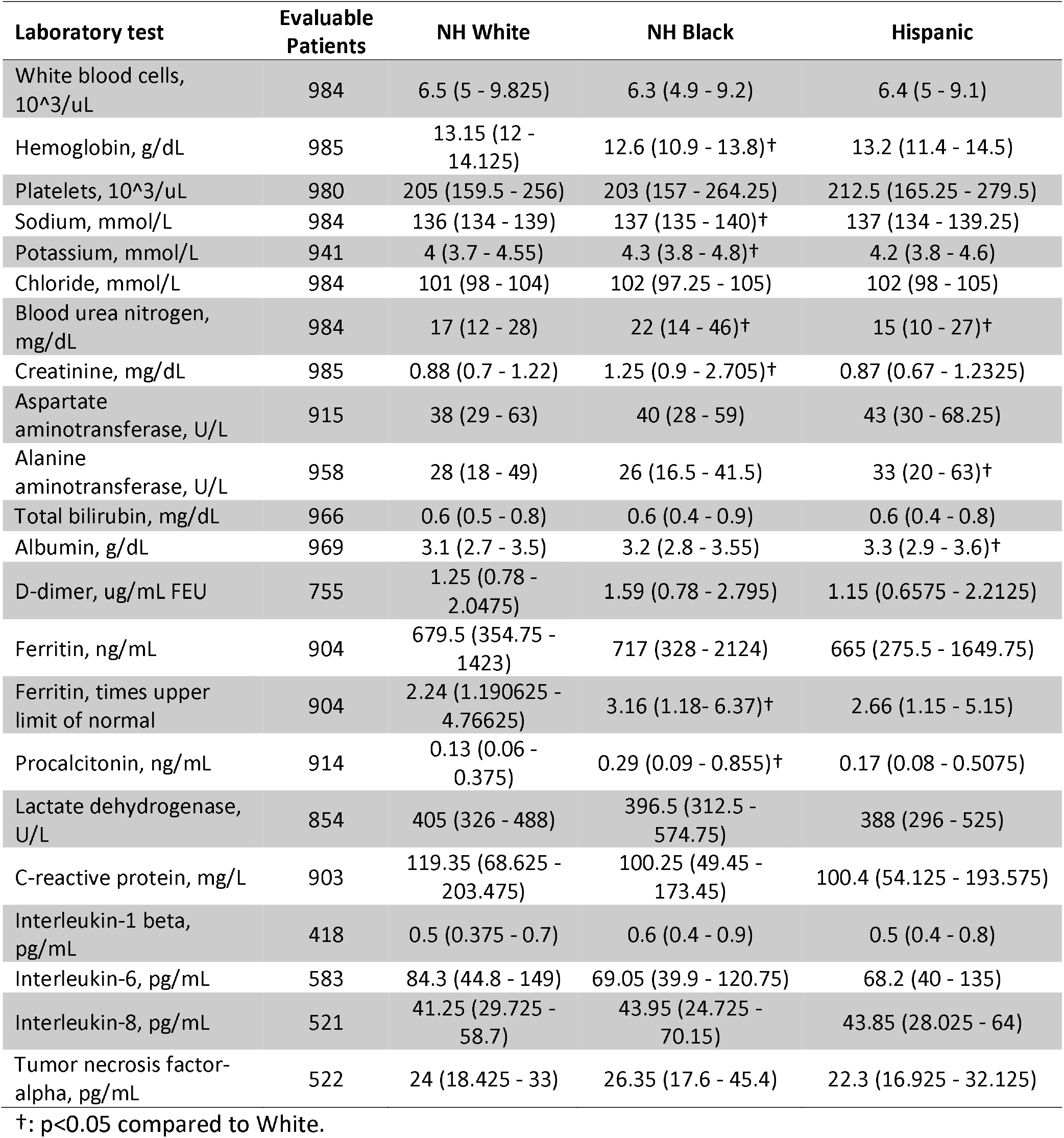
Baseline laboratory values among patients admitted to The Mount Sinai Hospital, by race/ethnicity

Unadjusted mortality rates were lower among Black (27.5% vs. 34.1%, p=0.006) and Hispanic (23.9% vs. 34.1%, p<0.001) patients compared to White patients. Rates of intensive care were not significantly different between Black and White (19.8% vs 21.5%, p=0.44) or Hispanic and White (23.4% vs 21.5%, p=0.36) patients.

### Baseline laboratory values

Among common lab values, Black patients had higher median baseline values of creatinine (1.25 vs. 0.88 mg/dL, p<0.001) and blood urea nitrogen (BUN, 22 vs. 17 mg/dL, p<0.001) than White patients, and Hispanic patients had higher baseline alanine aminotransferase values (ALT, 33 vs. 28 U/L, p=0.03) than White patients, consistent with the increased prevalence of chronic kidney disease among Black patients, and the increased prevalence of chronic liver disease among Hispanic patients.

Among inflammatory lab markers, Black patients had higher initial levels of procalcitonin (0.29 vs. 0.13 ng/mL, p<0.001), and more abnormal ferritin levels (3.16 vs. 2.24 times upper limit of normal, p=0.03) compared to White patients. There were no significant differences in baseline d-dimer, lactate dehydrogenase (LDH), C-reactive protein (CRP), interleukin-1-beta (IL-1B), interleukin-6 (IL-6), interleukin-8 (IL-8), or tumor necrosis factor-alpha (TNFa) levels.

### Race and ethnicity-specific factors associated with death

Using univariable and multivariable logistic regression, we evaluated the association of demographic, clinical, and laboratory variables with in-hospital mortality (**Table 3, Supplemental Table 2 & 3**).

**Table 3:** Multivariable model predicting in-hospital mortality, stratified by race/ethnicity

| Variable | White OR (95% CI) | Black OR (95% CI) | Hispanic OR (95% CI) | All patients OR (95% CI) |
| --- | --- | --- | --- | --- |
| Male | 1.51 (1.03-2.22) | 1.55 (1.09-2.19) | 1.04 (0.73-1.49) | 1.29 (1.05-1.58) |
| Age (yrs) | 1.07 (1.05-1.09) | 1.06 (1.04-1.07) | 1.05 (1.04-1.07) | 1.06 (1.05-1.07) |
| Race: NH White | Reference | Reference | Reference | Reference |
| Race: NH Black | NA | NA | NA | 1.03 (0.80-1.32) |
| Race: Hispanic | NA | NA | NA | 0.94 (0.73-1.21) |
| Manhattan facility | 0.45 (0.31-0.64) | 0.53 (0.37-0.75) | 0.62 (0.43-0.89) | 0.53 (0.43-0.65) |
| Hypertension | 0.86 (0.56-1.33) | 0.69 (0.45-1.06) | 0.78 (0.51-1.2) | 0.77 (0.61-0.98) |
| Diabetes | 0.99 (0.61-1.63) | 1.67 (1.09-2.58) | 1.21 (0.79-1.83) | 1.26 (0.98-1.63) |
| Coronary artery disease | 0.98 (0.58-1.65) | 1.14 (0.67-1.93) | 0.10 (0.57-1.73) | 1.03 (0.76-1.39) |
| Heart failure | 0.94 (0.48-1.85) | 1.11 (0.6-2.06) | 1.14 (0.55-2.36) | 1.05 (0.72-1.54) |
| Atrial fibrillation | 1.15 (0.66-2.02) | 1.27 (0.62-2.59) | 1.99 (0.97-4.11) | 1.38 (0.95-2.00) |
| Chronic kidney disease | 2.25 (1.18-4.28) | 1.71 (1.06-2.77) | 1.38 (0.80-2.38) | 1.70 (1.25-2.31) |
| COPD/asthma | 0.9 (0.48-1.68) | 0.80 (0.42-1.54) | 0.58 (0.32-1.06) | 0.75 (0.52-1.07) |
| Obesity | 0.72 (0.33-1.60) | 1.33 (0.69-2.58) | 1.59 (0.86-2.91) | 1.25 (0.86-1.83) |
| Cancer | 0.95 (0.49-1.85) | 1.15 (0.61-2.16) | 1.65 (0.88-3.07) | 1.25 (0.86-1.8) |
| Oxygen sat. <92% | 2.377 (1.6-3.54) | 2.51 (1.69-3.75) | 3.22 (2.24-4.61) | 2.71 (2.17-3.36) |

In univariate analysis, Black race (OR 0.73, 95% CI 0.59-0.91) and Hispanic ethnicity (OR 0.61, 95% CI 0.49-0.76) were associated with lower mortality compared to White race. However, race/ethnicity was not an independent predictor of mortality after adjusting for age, sex, comorbidities, and baseline hypoxia in this cohort (Black HR 1.03, 95% CI 0.80-1.32; Hispanic HR 0.94, 95% CI 0.73-1.21) (**Figure 2A**).

**Figure 2:**
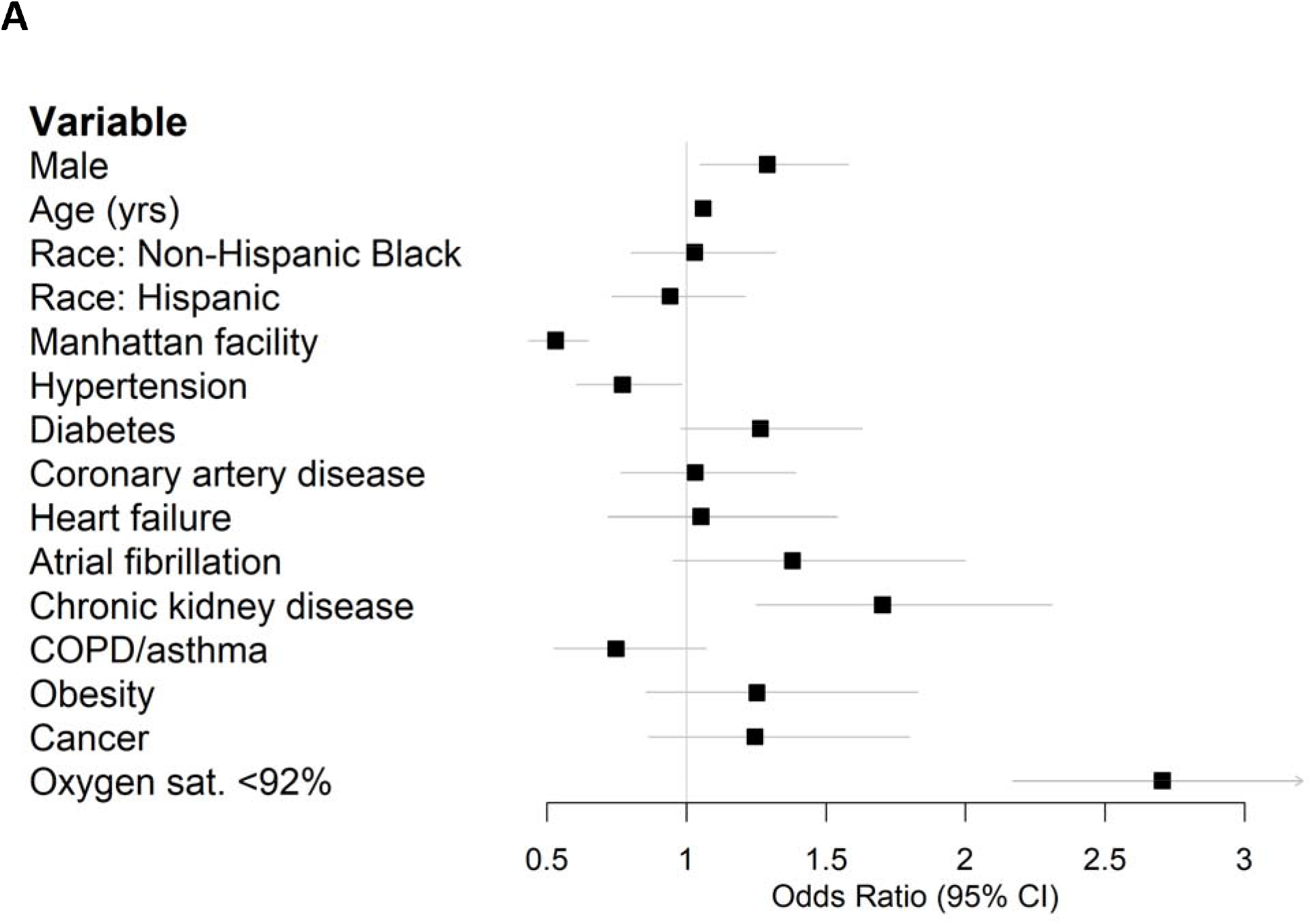
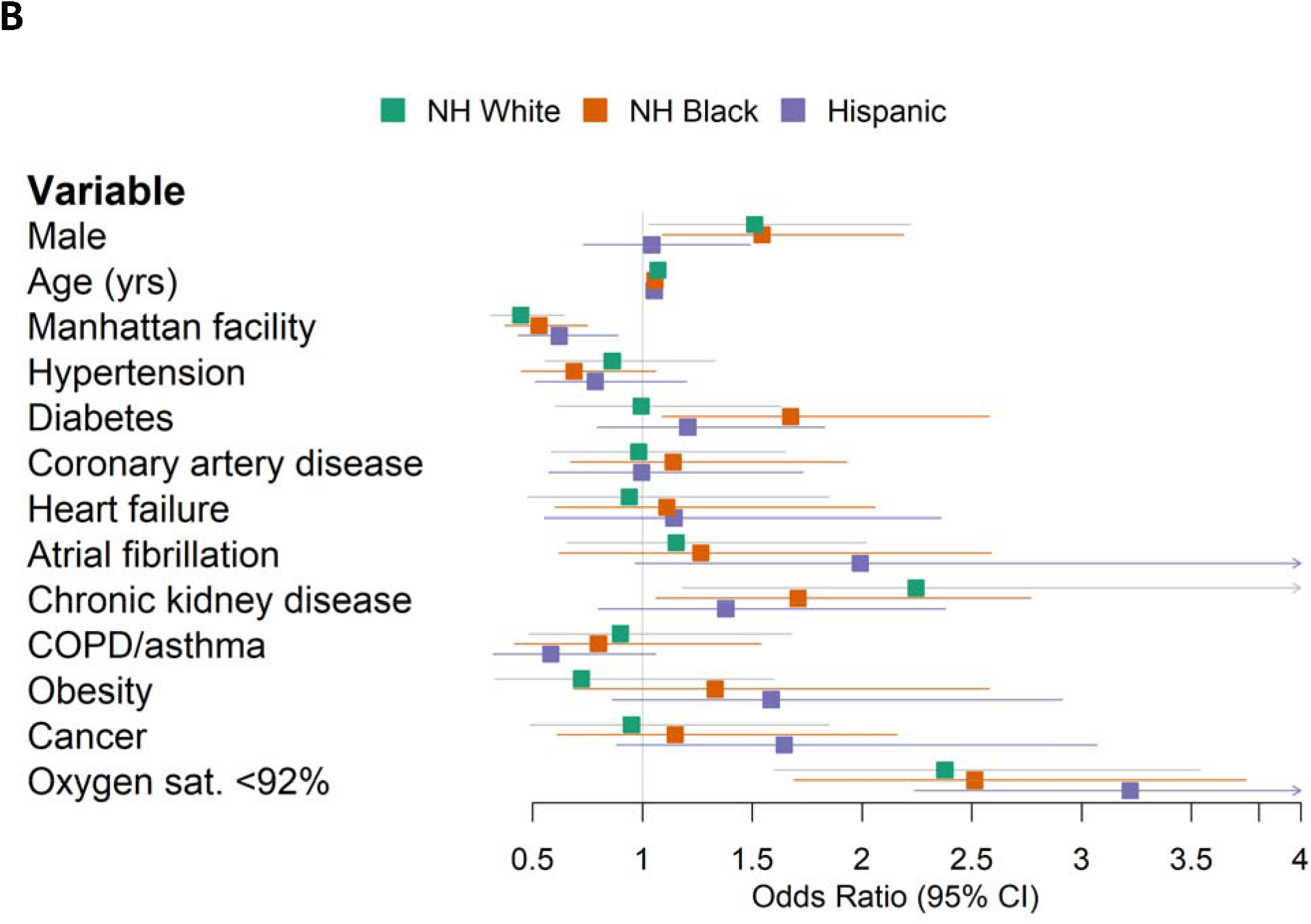
Forest plots of multivariable logistic regression results predicting in-hospital mortality. (A) All patients. (B) Stratified by race/ethnicity.

In multivariate models including interaction terms, there were suggestive but non-significant interactions between diabetes and Black race (p=0.09), as well as obesity and Black race (p=0.10). Diabetes was associated with an odds ratio of 1.67 (95% CI 1.09-2.58) in the Black-only model, and an odds ratio of 0.99 (95% CI 0.61-1.63) in the White-only model (**Figure 2B**). Obesity was associated with an odds ratio of 1.33 (95% CI 0.69-2.58) in the Black-only model, compared to an odds ratio of 0.72 (95% CI 0.33-1.60) in the White-only model (**Figure 2B**). Increased age and baseline hypoxia were consistently associated with increased mortality across all three groups (**Supplemental Table 4**).

Among inflammatory lab markers, univariate analysis showed significant associations with mortality for albumin (OR 0.624, 95% CI 0.52-0.75), CRP (OR 1.52, 95% CI 1.28-1.81), white blood cells (WBC, OR 1.48, 95% CI 1.22-1.79), IL-6 (OR 1.57, 95% CI 1.23-2.00), IL-8 (OR 4.31, 95% CI 1.76-10.5), LDH (OR 1.38, 95% CI 1.13-1.7), d-dimer (OR 1.33, 95% CI 1.1-1.62), and procalcitonin (OR 1.31, 95% CI 1.07-1.61) (**Supplemental Table 3**). After adjusting for age, sex, and hypoxia, CRP (OR 1.39, 95% CI 1.16-1.68), albumin (OR 0.75, 95% CI 0.61-0.91), IL-6 (OR 1.43, 95% CI 1.12-1.82), WBC (OR 1.35, 95% CI 1.08-1.65), and LDH (OR 1.34, 95% CI 1.07-1.68) remained independent predictors of mortality (**Supplemental Table 5**).

In multivariate models that included interaction terms, there was a significant interaction between Black race and IL-1B (p=0.04); elevated levels of IL-1B were associated with a higher risk of mortality in Black (OR 2.35, 95% CI 1.13-4.86) compared to White patients (OR 0.78, 95% CI 0.41-1.51) (**Figure 3, Supplemental Table 5**).

**Figure 3:**
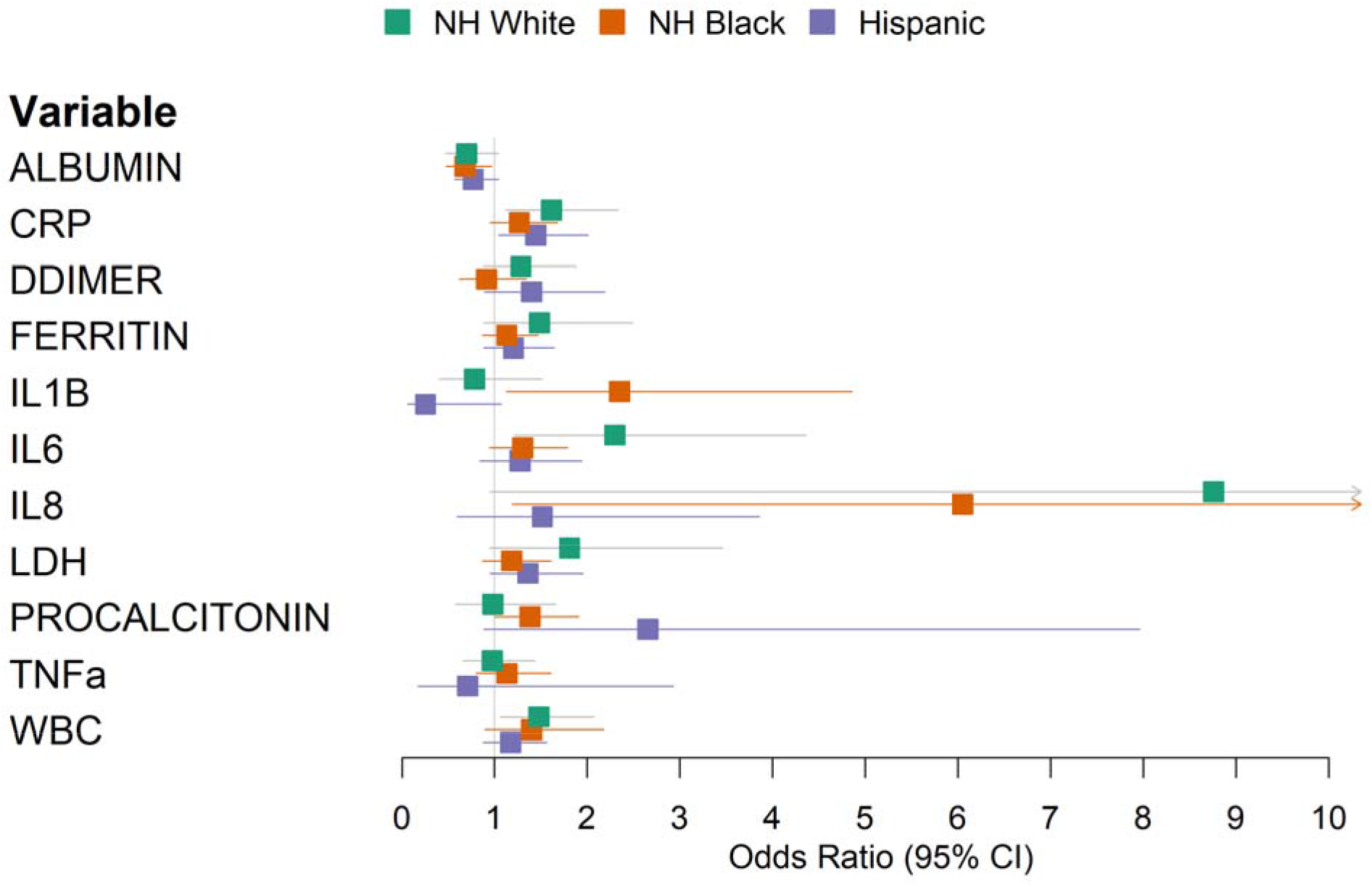
Forest plot of multivariable logistic regression results predicting in-hospital mortality using laboratory values, stratified by race/ethnicity. Models were adjusted for age, sex, and baseline hypoxia.

There were suggestive but non-significant interactions between Hispanic ethnicity and procalcitonin (p=0.07) and IL-8 (p=0.09) (**Supplemental Table 4**). Increased procalcitonin levels were associated with an odds ratio of 2.65 (95% CI 0.88-7.96) in Hispanic patients, compared to an odds ratio of 0.98 (95% CI 0.58-1.66) among White patients. Increased IL-8 levels were associated with an odds ratio of 1.51 (95% CI 0.59-3.86) among Hispanic patients, and an odds ratio of 8.76 (95% CI 0.95-80.7) among White patients (**Supplemental Table 5**).

## Discussion

Racial disparities in COVID-19 infections and outcomes have become apparent in both the US and elsewhere.^1–4^ The causes of these disparities are complex and multifactorial, involving social determinants of health as well as clinical and biological factors.^4,14^

In this study, set in New York City during the height of the initial COVID-19 surge, we describe the characteristics and outcomes of a diverse cohort of 2,406 White, Black, and Hispanic patients. The three groups differed significantly in demographic and clinical factors. White patients were older and showed higher rates of cardiovascular disease such as coronary artery disease atrial fibrillation. Black and Hispanic patients were younger, and had different comorbidity profiles, e.g. hypertension, diabetes, chronic kidney disease, and chronic liver disease.

Unadjusted in-hospital mortality was highest in White patients, but multivariable analysis showed that race/ethnicity was not an independent predictor of mortality in this cohort. It remains unclear whether race and ethnicity are independent risk factors for COVID-19 mortality after adjusting for confounding factors. Large national-level studies in the UK and Brazil have reported race as an independent predictor of mortality,^3,4^ whereas smaller studies in the US have not,^1,15,16^ possibly due to statistical power or population differences. In this New York City patient population, race/ethnicity was not an independent predictor for mortality.

In addition to describing this cohort, we aimed to test established COVID-19 risk factors for race/ethnicity-specific effects. Despite recapitulating several known risk factors, such as age, male sex, and hypoxia, we found only suggestive but non-significant interactions between Black race, diabetes and obesity, both of which tended to increase the mortality risk of Black patients to a greater degree than White patients. Notably, when analyzing inflammatory markers for their association with mortality, we found a significant interaction between Black race and the inflammatory cytokine IL-1B.

Excessive inflammation has emerged as an important aspect of COVID-19 pathophysiology, and the anti-inflammatory steroid dexamethasone has been shown to improve outcomes among those with severe disease.^17^ The interaction between Black race and IL-1B raises the possibility that differences in immunity may contribute to worse outcomes in some patients. Black Americans are at higher risk of autoimmune conditions such as systemic lupus erythematosus and lupus nephritis compared to White Americans, differences which can be linked in some cases to specific polymorphisms which are more common in African Americans.^18–21^ In our cohort, Black patients had higher levels of procalcitonin and ferritin at baseline compared to White patients, but other inflammatory markers (including IL-1B) were not significantly different. More attention should be paid in future studies to inflammatory profiles at baseline and over the course of infection, which may differ by race/ethnicity.

Our study has limitations that warrant specific mention. The dataset was derived from the electronic health record database without manual review, which may limit the completeness of comorbidity labels. Race and ethnicity were self-reported and were missing or unspecified in 17.4% of the initial cohort.

The strengths of our database include its size and the inclusion of 37.1% Hispanic patients, a vulnerable population in this pandemic which, to our knowledge, has not been specifically examined in the literature to date. Additionally, our near-complete follow-up of the cohort’s hospital outcomes (99.3%) strengthens the validity of our findings.

In conclusion, our analysis of a diverse cohort drawn from the New York metropolitan area highlights both similarities and important differences across racial and ethnic groups in risk factors for death among hospitalized COVID-19 patients.

## Data Availability

Please contact the authors for data requests.

## Acknowledgment

We dedicate this work to the frontline health workers and staff of the Mount Sinai Healthcare System. This work was supported in part through the Mount Sinai Data Warehouse (MSDW) resources and staff expertise provided by Scientific Computing at the Icahn School of Medicine at Mount Sinai.

## Supplemental Materials

*Supplemental Table 1: Self-reported ethnicities which were classified as (A) Hispanic, and (B) Non-Hispanic Black*.

*Supplemental Table 2: Univariable logistic regression using demographic and clinical factors to predict in-hospital mortality, stratified by race/ethnicity*.

*Supplemental Table 3: Univariable logistic regression using standardized laboratory values to predict in-hospital mortality, stratified by race/ethnicity*.

*Supplemental Table 4: Interaction p-values from multivariable logistic regression including interaction terms between race/ethnicity and demographic, clinical, or laboratory factors*.

*Supplemental Table 5: Multivariable logistic regression using standardized laboratory values to predict in-hospital mortality, stratified by race/ethnicity, adjusting for age, sex, and baseline hypoxia*.

## Notes

### Competing Interest Statement

The authors have declared no competing interest.

